# Multiomics identifies unique modulators of calf muscle pathophysiology in peripheral artery disease and chronic kidney disease

**DOI:** 10.1101/2024.09.30.24314668

**Authors:** Kyoungrae Kim, Trace Thome, Caroline Pass, Lauren Stone, Nicholas Vugman, Victoria Palzkill, Qingping Yang, Kerri A. O’Malley, Erik M. Anderson, Brian Fazzone, Feng Yue, Scott A. Berceli, Salvatore T. Scali, Terence E. Ryan

**Affiliations:** Department of Applied Physiology and Kinesiology, The University of Florida, Gainesville, FL, USA; Department of Surgery, The University of Florida, Gainesville, FL, USA; Department of Animal Sciences, The University of Florida, Gainesville, FL, USA; Center for Exercise Science, The University of Florida, Gainesville, FL, USA; Myology Institute, The University of Florida, Gainesville, FL, USA

**Keywords:** peripheral artery disease, renal, uremia, mitochondria, muscle

## Abstract

**BACKGROUND:** Chronic kidney disease (CKD) has emerged as a significant risk factor that accelerates atherosclerosis, decreases muscle function, and increases the risk of amputation or death in patients with peripheral artery disease (PAD). However, the modulators underlying this exacerbated pathobiology are ill-defined. Recent work has demonstrated that uremic toxins are associated with limb amputation in PAD and have pathological effects in both the limb muscle and vasculature. Herein, we utilize multiomics to identify novel modulators of disease pathobiology in patients with PAD and CKD.

**METHODS:** A cross-sectional study enrolled four groups of participants: controls without PAD or CKD (n=28), patients with PAD only (n=46), patients with CKD only (n=31), and patients with both PAD and CKD (n=18). Both targeted (uremic toxins) and non-targeted metabolomics in plasma were performed using mass spectrometry. Calf muscle biopsies were used to measure histopathology, perform bulk and single-nucleus RNA sequencing (snRNAseq), and assess mitochondrial function. Differential gene and metabolite analyses, as well as pathway and gene set enrichment analyses were performed.

**RESULTS:** Patients with both PAD and CKD exhibited significantly lower calf muscle strength and smaller muscle fiber areas compared to controls and those with only PAD. Compared to controls, mitochondrial function was impaired in CKD patients, with or without PAD, but not in PAD patients without CKD. Plasma metabolomics revealed substantial alterations in the metabolome of patients with CKD, with significant correlations observed between uremic toxins (e.g., kynurenine, indoxyl sulfate) and both muscle strength and mitochondrial function. RNA sequencing analyses identified downregulation of mitochondrial genes and pathways associated with protein translation in patients with both PAD and CKD. Single nucleus RNA sequencing further highlighted a mitochondrial deficiency in muscle fibers along with unique remodeling of fibro-adipogenic progenitor cells in patients with both PAD and CKD, with an increase in adipogenic cell populations.

**CONCLUSIONS:** CKD significantly exacerbates ischemic muscle pathology in PAD, as evidenced by diminished muscle strength, reduced mitochondrial function, and altered transcriptome profiles. The correlation between uremic toxins and muscle dysfunction suggests that targeting these metabolites may offer therapeutic potential for improving muscle health in PAD patients with CKD.

## INTRODUCTION

Peripheral arterial disease (PAD) is a condition characterized by atherosclerosis in the blood vessels of the lower extremities, affecting approximately 8-12 million Americans, and is a leading cause of cardiovascular mortality^1,2^. Common risk factors for PAD include age, smoking, diabetes, hypertension, and dyslipidemia. Notably, chronic kidney disease (CKD) is also strongly associated with PAD, contributing to the development of atherosclerosis, decreased muscle function, and an increased risk of amputation or death in patients with PAD^3–9^. Furthermore, CKD is linked to higher failure rates of both endovascular and open surgical revascularization procedures, which are commonly used in the management of PAD^3,10^. Recent work has reported a strong correlation between uremic metabolites— accumulating in CKD—and the risk of adverse limb events in patients with PAD^11^. Both PAD and CKD have been associated with skeletal muscle myopathy^12–24^, suggesting that muscle tissue may be a key site where these two diseases intersect. Importantly, muscle function and exercise performance have been established as strong predictors of morbidity and mortality in patients with PAD^13,15,25–27^. Despite these associations, the mechanisms underlying the exacerbated skeletal myopathy in patients with both PAD and CKD remain poorly understood. A deeper analysis could reveal novel targets for preventing or ameliorating muscle pathology in these patients. This cross-sectional study aimed to characterize the impact of PAD and CKD on skeletal muscle pathology by comparing calf muscle strength, mitochondrial function, and histopathology in four groups: 1) PAD patients, 2) PAD with CKD patients, 3) CKD patients, and 4) non-PAD and non-CKD controls. A multiomic approach, including bulk and single nucleus RNA sequencing of calf muscle specimens and both targeted and untargeted plasma metabolomics, was employed to identify novel regulators of skeletal muscle pathology

## MATERIALS AND METHODS

Detailed descriptions of the materials and experimental procedures can be found in the Expanded Material and Methods section of the **Supplemental Material**. The data supporting the conclusions of this study are available from the corresponding author. A Major Resources Table is available in the **Supplemental Material**.

### Study Population

This was a cross-sectional study involving participants without PAD or CKD, those with PAD only, those with CKD only, and those with both PAD and CKD. Participants were recruited through UF Health and the Malcom Randall VA Medical Centers in Gainesville, FL. This study was approved by the institutional review boards at the University of Florida and the Malcom Randall VA Medical Center (Protocol IRB201801553). All study procedures were carried out according to the Declaration of Helsinki and participants were fully informed about the research and informed consent was obtained. Patients with a normal ABI (greater than 0.9) or those with an abnormal ABI (less than 0.9) indicating a diagnosis of PAD with a Rutherford Stage between 3-5 set as inclusion criteria. Patients with non-atherosclerotic occlusive disease were excluded (acute limb ischemia, vasculitis, Buerger’s disease, etc.). Inclusion criteria for CKD included an estimated glomerular filtration rate (eGFR) between 15 and 50 mL/min/1.73m^2^ for greater than three months. Medical history, race, smoking history, and demographics were obtained by self-report. Co-existing medical conditions and medication usage was obtained through review of medical records. Non-PAD controls were recruited within the University of Florida Hospital System and Malcom Randall VA medical center and were free from peripheral vascular disease.

### Calf muscle testing

A detailed description of analysis of calf muscle strength, muscle biopsy technique, mitochondrial function, muscle histopathology, and bulk and single nucleus RNA sequencing can be found on the **Supplemental Material**.

### Plasma Metabolomics

A blood sample was obtained from a peripheral antebrachial vein using EDTA tubes and was subsequently centrifuged and plasma was frozen at −80°C until analysis. Sample underwent untargeted metabolomic analysis (Global Discovery platform) at a core laboratory using a well-validated Ultrahigh Performance Liquid Chromatography-Tandem Mass Spectroscopy (UPLC-MS/MS) platform (Metabolon, Inc.; Morrisville, NC, USA). The platform combines multiple sample preprocessing protocols and a comprehensive reference library, resulting in quantitative estimates of metabolite abundance covering a broad spectrum of chemical classes, including amino acids, carbohydrates, lipids, nucleotides, peptides and vitamins, xenobiotic substances, and pharmaceutical and food preservative compounds. A detailed description of the analytical protocol, metabolite identification, and normalization procedures is included in the Supplementary Methods. Samples were randomized across the platform run with quality control samples spaced evenly among the injections. Metabolite abundance was estimated from the area under the curve for annotated peaks in the mass spectrogram (mass spectral counts) and was normalized for batch and run day. Samples were also processed for targeted metabolomic analysis of kynurenine, tryptophan, 3-indoxyl sulfate, p-cresol sulfate (pCS), 4-ethylphenyl sulfate (4-EPS), 3-indolelactic acid, and 3-indolepropionic acid in which quantitative analysis was performed using standard curves for each metabolite.

### Statistical analysis

Normality of data was tested with the Shapiro-Wilk test and inspection of QQ plots. Data involving comparisons of normally distributed data from groups were analyzed using a one-way ANOVA. Data involving comparisons of non-normally distributed data from groups were analyzed using a Kruskal-Wallis test. When appropriate, Tukey’s posthoc testing was used to assess differences between groups. To assess relationships between two variables, Pearson correlation analysis was employed. Chi-Square analysis was used to determine differences in population proportions for relevant clinical characteristics. For bulk and snRNAseq, the Benjamini–Hochberg method was used to calculate false discovery rate corrected *P-*values. In all cases, *P* < 0.05 was considered statistically significant. All statistical testing was conducted using R-studio, Python, or GraphPad Prism software (version 9.0). Data are presented as the mean ± standard deviation unless otherwise specified.

## RESULTS

### Patients with PAD and CKD have significantly lower calf muscle strength and myofiber areas

Fourty-six patients with PAD with normal kidney function, 18 patients with PAD and CKD, 31 patients with CKD only (no PAD), and 28 people with PAD or CKD were included in this study (N=123 total). The participant characteristics are shown in **Table 1**. Calf muscle (plantar flexors) strength was significantly lower in patients with PAD (*P*=0.0409), patients with CKD (*P*=0.0193), and patients with PAD and CKD (*P*<0.0001) when compared with controls that were free from PAD and CKD (**Figure 1A**). PAD patients with CKD had significantly lower calf muscle strength than PAD patients with normal kidney function (*P*=0.0103). After normalizing calf muscle strength to body weight, significant differences remained for PAD vs. PAD+CKD and control vs. PAD+CKD comparisons. Analysis of the mean myofiber cross sectional area demonstrated that PAD, CKD, and PAD+CKD groups had significantly smaller Type I and Type II myofibers compared to control participants (**Figure 1B,C**), however, the relative proportion fiber types was not significantly different (**Figure 1D**). Within the cohort, a significant correlation (Pearson r = 0.3160, *P*=0.0390) was observed between calf muscle strength and the mean myofiber area (**Figure 1E**). These findings reveal a clear deterioration in muscle function in PAD patients with CKD.

**Figure 1.**
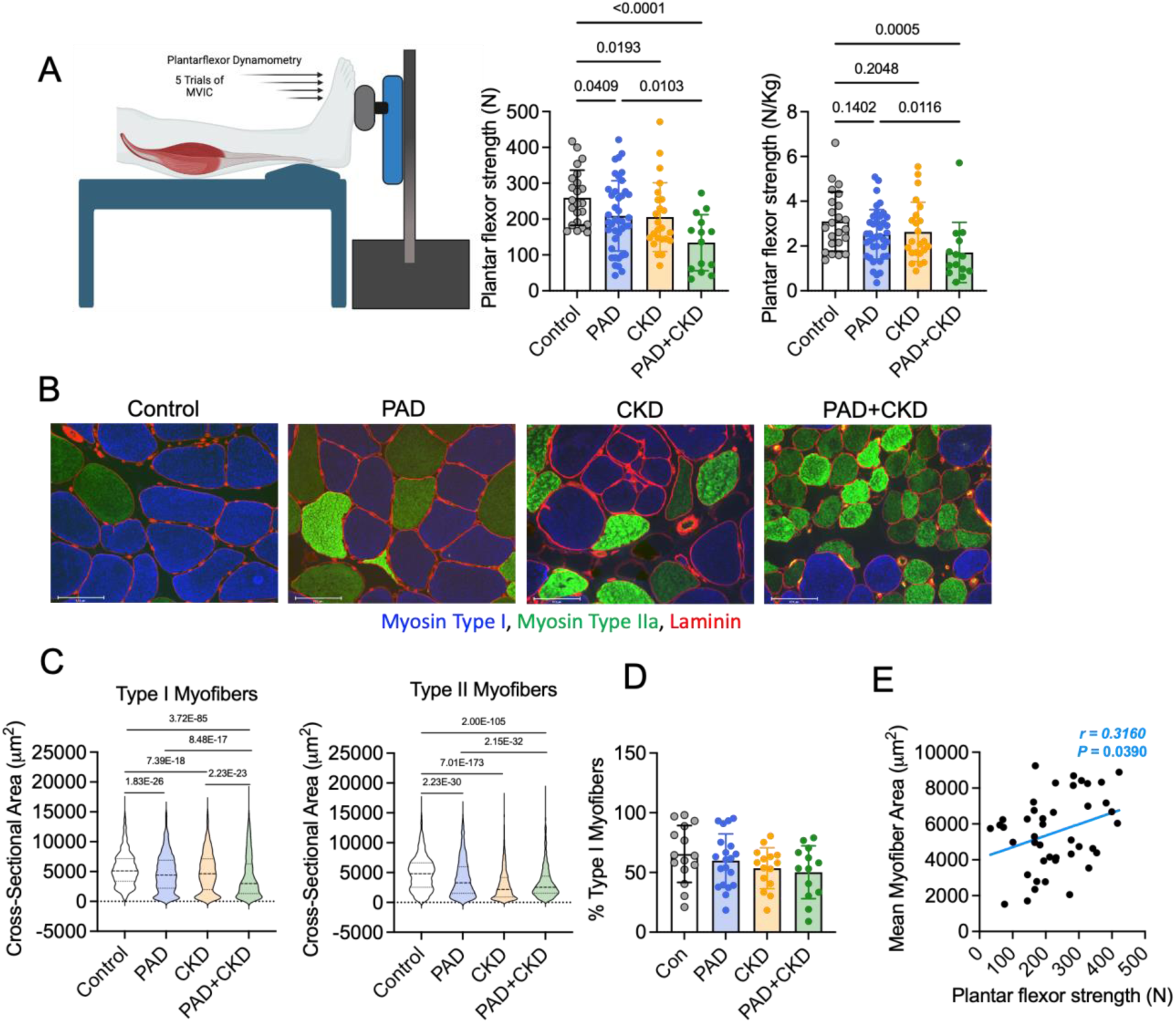
Calf muscle strength and muscle fiber area is lowest in patients with PAD and CKD. (**A**) Calf muscle strength in absolute and normalized to body weight (n=102). (**B**) Representative immunofluorescence images of gastrocnemius muscle biopsies with fiber types and sizes labeled. (**C**) Violin plots showing the cross-sectional areas of Type I (n=8816 myofibers) and Type II (n=8395) myofibers in each patient group (n=63 patients). (**D**) Quantification of the percentage of Type I fibers (n=63 patients). (E) Pearson correlation between calf muscle strength and mean myofiber area (both Type I and II myofibers) (n=43 patients). Panel A was analyzed using a Kruskal-Wallis Test with Dunn’s test for pairwise comparisons. Panel C and D were analyzed using a one-way ANOVA with Tukey’s post hoc testing. Error bars represent the standard deviation.

**TABLE 1:** Participant Characteristics.

| | Control<br>(N=28) | PAD (N=46) | CKD<br>(N=31) | PAD+CKD<br>(N=18) | *P* value<br>($\chi^2$ test or<br>One-way<br>ANOVA) |
| --- | --- | --- | --- | --- | --- |
| Physical Characteristics |  |  |  |  |  |
| Mean age, y (SD) | 64.4 (12.0) | 64.5 (8.6) | 62.1 (12.4) | 65.5 (9.0) | 0.6869 |
| Female sex, n (%) | 8 (28.6) | 18 (39.1) | 13 (41.9) | 9 (50.0) | 0.5111 |
| Non-White, n (%) | 4 (14.3) | 10 (21.7) | 12 (38.7) | 6 (33.3) | 0.1366 |
| Non-Hispanic, n (%) | 26 (92.9) | 40 (87.0) | 31 (100.0) | 18 (100.0) | 0.0835 |
| BMI, kg/m<sup>2</sup> (SD) | 29.1 (6.3) | 27.6 (6.0) | 30.0 (6.9) | 28.3 (6.5) | 0.4777 |
| Disease Characteristics |  |  |  |  |  |
| ABI (SD) | 1.00 (0.08) | 0.60 (0.26) | 1.04 (0.16) | 0.65 (0.24) | <0.0001 |
| TBI (SD) | 0.81 (0.17) | 0.36 (0.21) | 0.75 (0.15) | 0.34 (0.23) | <0.0001 |
| BUN, (mg/dL) (SD) | 15.8 (5.8) | 14.6 (5.2) | 35.0 (18.0) | 38.8 (25.6) | <0.0001 |
| Creatinine, (mg/dL) (SD) | 0.9 (0.2) | 0.9 (0.2) | 4.4 (2.5) | 3.9 (3.7) | <0.0001 |
| eGFR, (mL/min/1.73m<sup>2</sup>) (SD) | 73.9 (19.3) | 70.2 (17.0) | 26.0 (12.2) | 30.1 (16.2) | <0.0001 |
| Tobacco use |  |  |  |  |  |
| Former smoker, n (%) | 18 (66.7) | 44 (95.7) | 9 (30.0) | 14 (77.8) | <0.0001 |
| Current smoker, n (%) | 7 (25.9) | 13 (28.3) | 1 (3.3) | 6 (33.3) | 0.0303 |
| Medical History |  |  |  |  |  |
| Diabetes type I or II, n (%) | 9 (32.1) | 20 (43.5) | 16 (51.6) | 14 (77.8) | 0.0208 |
| Hypertension, n (%) | 18 (64.3) | 38 (82.6) | 30 (96.8) | 17 (94.4) | 0.0042 |
| Hyperlipidemia, n (%) | 17 (60.7) | 35 (76.1) | 18 (58.1) | 15 (83.3) | 0.1416 |
| Coronary artery disease, n (%) | 10 (35.7) | 23 (50.0) | 7 (22.6) | 6 (33.3) | 0.1028 |
| Congestive heart failure, n (%) | 6 (21.4) | 9 (19.6) | 9 (29.0) | 7 (38.9) | 0.3890 |
| Medication used |  |  |  |  |  |
| Aspirin, n (%) | 15 (53.6) | 35 (76.1) | 13 (41.9) | 12 (66.7) | 0.0183 |
| ACE inhibitor, n (%) | 5 (17.9) | 17 (37.0) | 7 (22.6) | 4 (22.2) | 0.2581 |
| Angiotensin receptor blocker, n (%) | 4 (14.3) | 8 (17.4) | 8 (25.8) | 7 (38.9) | 0.1869 |
| Statin, n (%) | 21 (75.0) | 37 (80.4) | 22 (71.0) | 17 (94.4) | 0.2509 |
| Cilostazol, n (%) | 0 (0.0) | 14 (30.4) | 0 (0) | 4 (22.2) | 0.0006 |
| Beta Blocker, n (%) | 14 (50.0) | 25 (54.3) | 19 (61.3) | 15 (83.3) | 0.1201 |
| Anticoagulant, n (%) | 3 (10.7) | 17 (37.0) | 8 (25.8) | 8 (44.4) | 0.0430 |
| Antiplatelet, n (%) | 5 (17.9) | 12 (26.1) | 7 (22.6) | 2 (11.1) | 0.5764 |
| Abbreviations: BMI = body mass index; ABI = ankle-brachial index; TBI = toe-brachial index; BUN = blood urea nitrogen; eGFR = estimated glomerular filtration rate; ACE = angiotensin converting enzyme |  |  |  |  |  |

### Calf muscle mitochondrial function is negatively impacted by CKD, but not PAD alone

High-resolution respirometry and fluorescence spectroscopy were used to assess muscle mitochondrial function in freshly permeabilized muscle fiber bundles using a creatine kinase clamp to perform a ‘mitochondrial stress test” across a range of physiologically relevant energy demands (**Figure 2A**). Oxygen consumption (*J*O_2_) was lower in patients with CKD and PAD+CKD, but nearly identical in patients with only PAD and control participants (**Figure 2B**). Quantification of the slope of oxygen consumption vs. energy demand (ΔG_ATP_), termed OXPHOS conductance, represents the sensitivity of the mitochondrial system to respond to increases in energy demand. In this case, a higher OXPHOS conductance is interpreted as better mitochondrial function. OXPHOS conductance was not different between controls and PAD patients with normal kidney function (*P*=0.97), however CKD patients without PAD and PAD+CKD patients had significantly lower OXPHOS conductance compared to controls (*P*=0.0123 and *P*=0.0054 respectively). PAD+CKD patients had significantly lower OXPHOS conductance than PAD patients without CKD (**Figure 2C**). Citrate synthase activity, a biomarker of mitochondrial content in skeletal muscle, was not significantly lower in CKD (*P*=0.095) and PAD+CKD (*P*=0.63) groups compared to controls (**Figure 2D**). Mitochondrial H_2_O_2_ emission was not significantly different between groups across the levels of energy demand (**Figure 2E**) or without energy demand (**Figure 2F**) when fueled by pyruvate/malate/octanoylcarnitine. When fueled with succinate in the absence of energy demand with or without inhibitors of the matrix antioxidant systems, PAD+CKD muscles had lower mitochondrial H_2_O_2_ emission than controls (**Figure 2G**). Succinate dehydrogenase (SHD) enzyme activity assessed histochemically on thin sections of muscle showed a lower SDH activity in CKD and PAD+CKD groups (**Figure 2H**).

**Figure 2.**
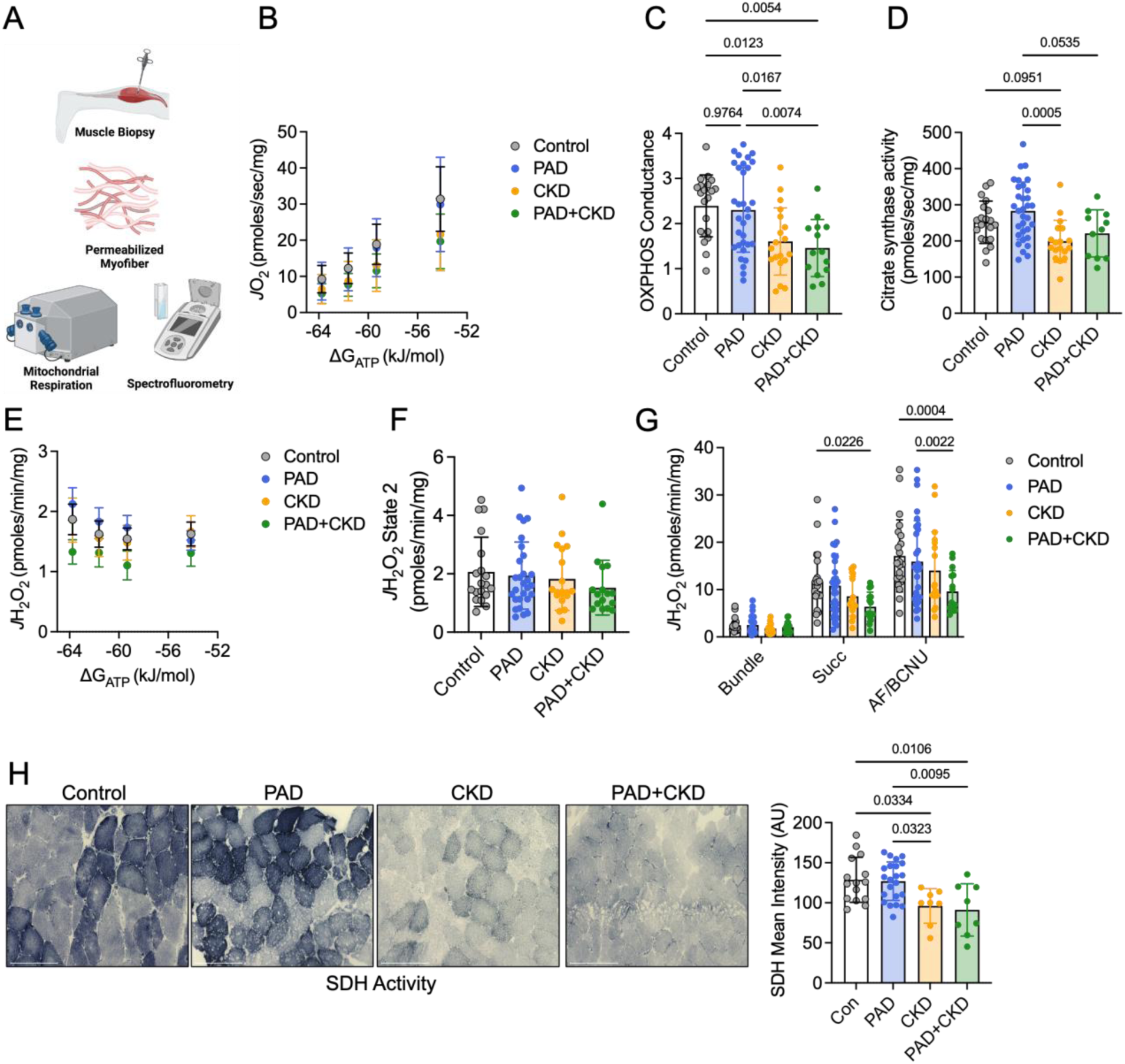
Skeletal muscle mitochondrial function is significantly impaired in patients with CKD and PAD+CKD. (**A**) Graphical depiction of mitochondrial analyses using permeabilized myofibers. (**B**) Relationship between oxygen consumption (*J*O_2_) and energy demand (ΔG_ATP_) when mitochondria were fueled with pyruvate, malate, and octanoylcarnitine. (**C**) Quantification of the conductance (the slope of *J*O_2_ and ΔG_ATP_ relationship) (n=86 patients). (**D**) Citrate synthase activity in gastrocnemius muscle (n= 81 patients). (**E**) Relationship between mitochondrial hydrogen peroxide production (*J*H_2_O_2_) and energy demand (ΔG_ATP_) when mitochondria were fueled with pyruvate, malate, and octanoylcarnitin. (**F**) Quantification of *J*H_2_O_2_ under state 2 (no energy demand) conditions (n=82 patients). (**G**) Quantification of *J*H_2_O_2_ with mitochondrial fueled with succinate followed by inhibition of the matrix antioxidant systems (with AF/BCNU) (n=85 patients). Analyzed via two-way ANOVA with Tukey’s post hoc. (**H**) Representative images and quantification of succinate dehydrogenase (SDH) activity in muscle sections (n= patients. Panels C, D, F, and H were analyzed with a one-way ANOVA with Tukey’s post hoc. Error bars represent the standard deviation.

### CKD significantly alters the plasma metabolome

Next, we performed targeted metabolomics to quantify the absolute abundance of known uremic toxins including 3-indoxyl sulfate, p-cresol sulfate (pCS), 4-ethylphenyl sulfate (4-EPS), 3-indolelactic acid, 3-indolepropionic acid, kynurenine (L-Kyn), tryptophan (Trp), and the L-Kyn/Trp ratio in plasma of this cohort. As expected, CKD and PAD+CKD groups had significantly higher levels of all these uremic toxins (**Figure 3A**). No differences were found between controls and patients with PAD only. We also performed global untargeted metabolomics on the cohort which resulted in identification of 1557 metabolites/biochemicals, 1277 of which were identified in the compound library and 277 with unknown identity (**Figure 3B**). Principal component analysis revealed two relatively distinct clusters which separate the patients with and without CKD (**Figure 3C**). Compared to controls, patients with PAD had relatively similar plasma metabolomes with 99 differentially abundant metabolites (46 up and 53 down). In contrast, patients with CKD had 763 differentially abundant metabolites compared to controls (623 up and 140 down). Consistent with these findings, PAD+CKD patients had 578 differentially abundant metabolites (521 up and 57 down) compared to PAD patients. A heatmap with hierarchical clustering for all patients and metabolites is shown in **Figure 3D**. A Venn diagram showing the shared and distinct numbers of differentially abundant metabolites across comparisons is shown in **Figure 3E**. Pathway analysis revealed three main classes of metabolites were impacted by disease classification including “Uremic Toxins”, “Tissue Remodeling”, and Microbiome-related metabolites” (**Supplemental Figure 1**). The complete metabolomics dataset and pathway analysis can be found in Supplemental Datasets 1 and 2 respectively.

**Figure 3.**
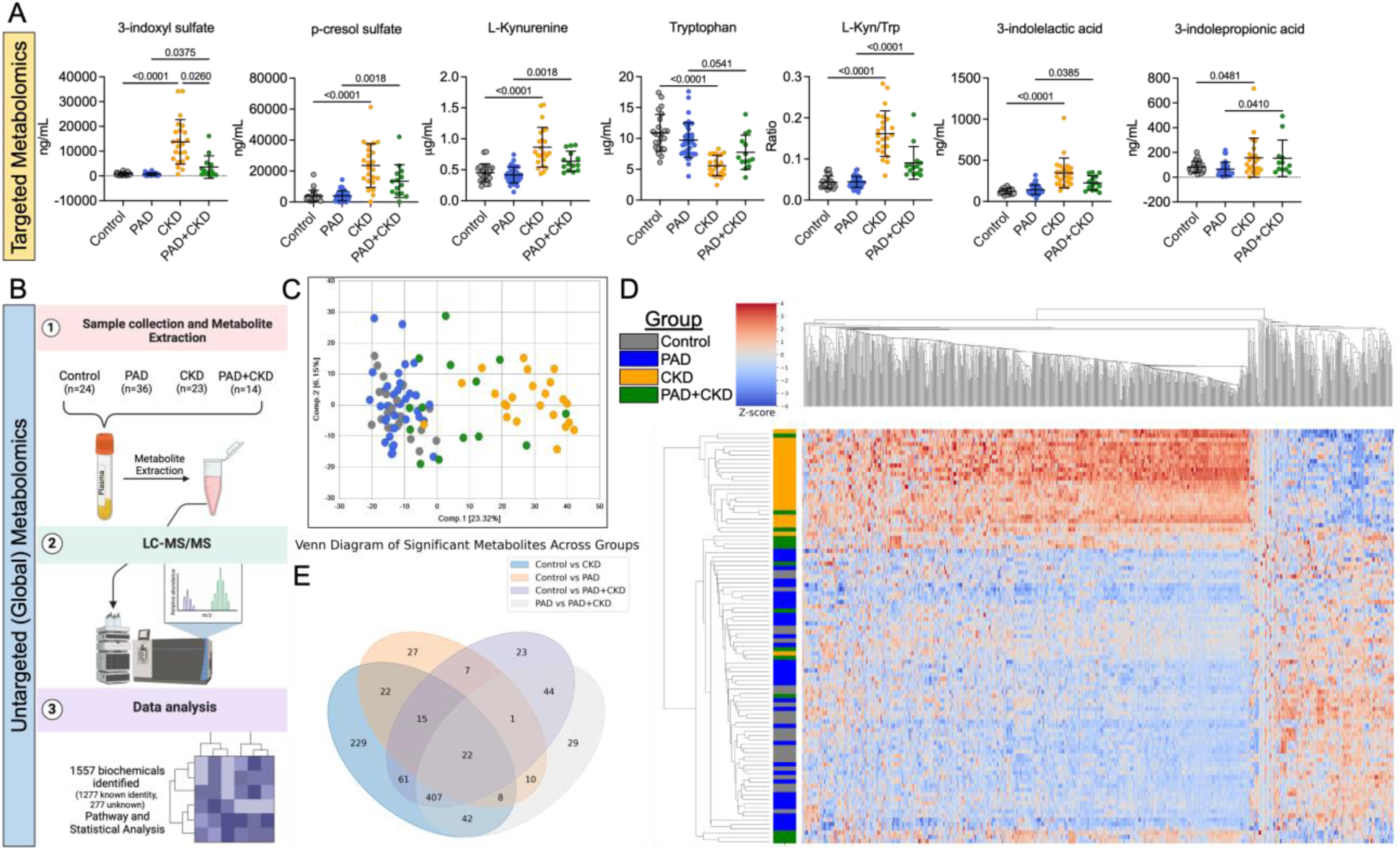
CKD profoundly impacts the plasma metabolome independent of PAD. (**A**) Targeted plasma metabolomic quantification of uremic toxins (n=97 patients). Analysis done using one-way ANOVA with Tukey’s post hoc. (**B**) Graphic of experimental design and overall metabolite detection for untargeted (global) metabolomics in patient plasma. (**C**) Principal component analysis demonstrates separation between CKD and non-CKD patients, independent of PAD. (**D**) A heatmap with hierarchical clustering of metabolites and patients. (**E**) Venn diagram showing significant metabolite differences across group comparisons. Error bars represent the standard deviation.

### Skeletal muscle mitochondrial function and calf muscle strength are negatively correlated with tryptophan-derived uremic toxins

Previous studies have reported that tryptophan-derived uremic toxins, such as indoxyl sulfate and kynurenine, were negatively associated with muscle health and mitochondrial function^23,28–31^. Thus, we perform Pearson correlation analyses to determine if mitochondrial function (OXPHOS conductance) or muscle strength were related to these uremic toxins. Strikingly, kynurenine, L-Kyn/Trp ratio, and indoxyl sulfate had significant inverse correlations with both mitochondrial function (**Figure 4A**) and calf muscle strength (**Figure 4B**) in this cohort.

**Figure 4.**
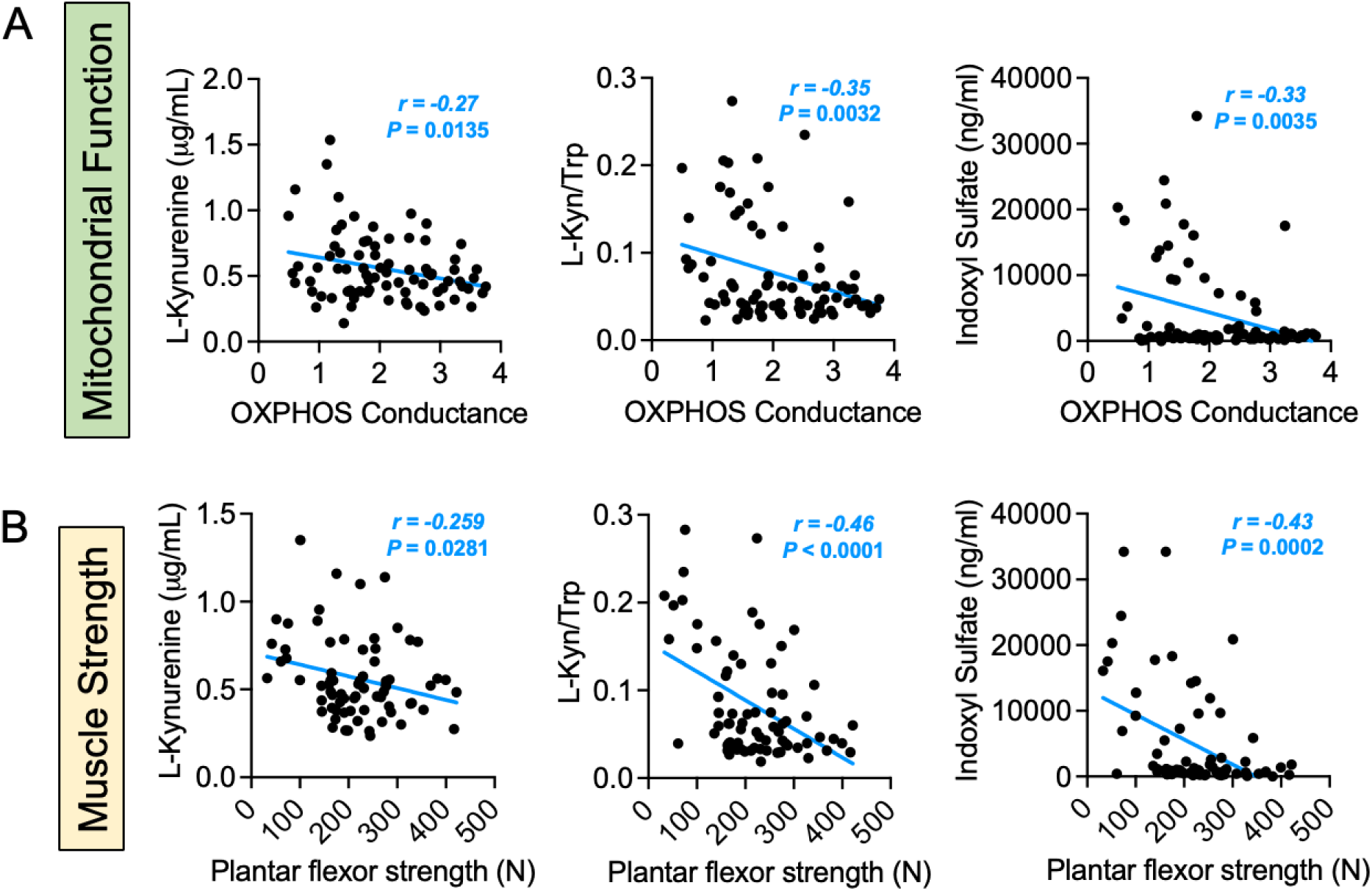
Uremic toxin levels have an inverse relationship with calf muscle mitochondrial function and strength. (**A**) Pearson correlations between OXPHOS conductance and L-Kynurenine, L-Kyn/Trp ratio, and indoxyl sulfate levels (n=81 patients). (**B**) Pearson correlations between calf muscle strength and L-Kynurenine, L-Kyn/Trp ratio, and indoxyl sulfate levels (n=72 patients). Statistical analyses performed using two-tailed Pearson correlation.

### Whole muscle and single nucleus RNA sequencing identifies transcriptome changes in PAD patients with CKD

To understand how CKD impacts the skeletal muscle transcriptome, we performed bulk RNA sequencing on total RNA isolated from a portion of the gastrocnemius muscle biopsy of 88 patients. Gene expression was quantified for 24,871 genes. Comparing PAD to controls, 52 differentially expressed genes were detected (12 downregulated and 40 upregulated) (**Supplemental Figure 2A-D**). Comparing CKD to controls, 602 differentially expressed genes were detected (300 downregulated and 302 upregulated) (**Supplemental Figure 2E-H**). Comparing patients with PAD to those with PAD+CKD, partial least squares discriminant analysis (PLS-DA) showed two distinct clusters (**Figure 5A**). Patients with PAD+CKD had 361 differentially expressed genes from patients with only PAD (189 downregulated and 172 upregulated) (**Figure 5B**). GSEA in this comparison revealed that down regulated genes were significantly enriched in pathways related to “cellular respiration” and “oxidative phosphorylation”, whereas significantly upregulated genes were enriched in pathways related to “cytoplasmic translation”, “ribosome biogenesis”, and “rRNA processing” (**Figure 5C**). Full results of the DEG and GSEA analysis for each comparison can be found in Supplemental Dataset 3.

**Figure 5.**
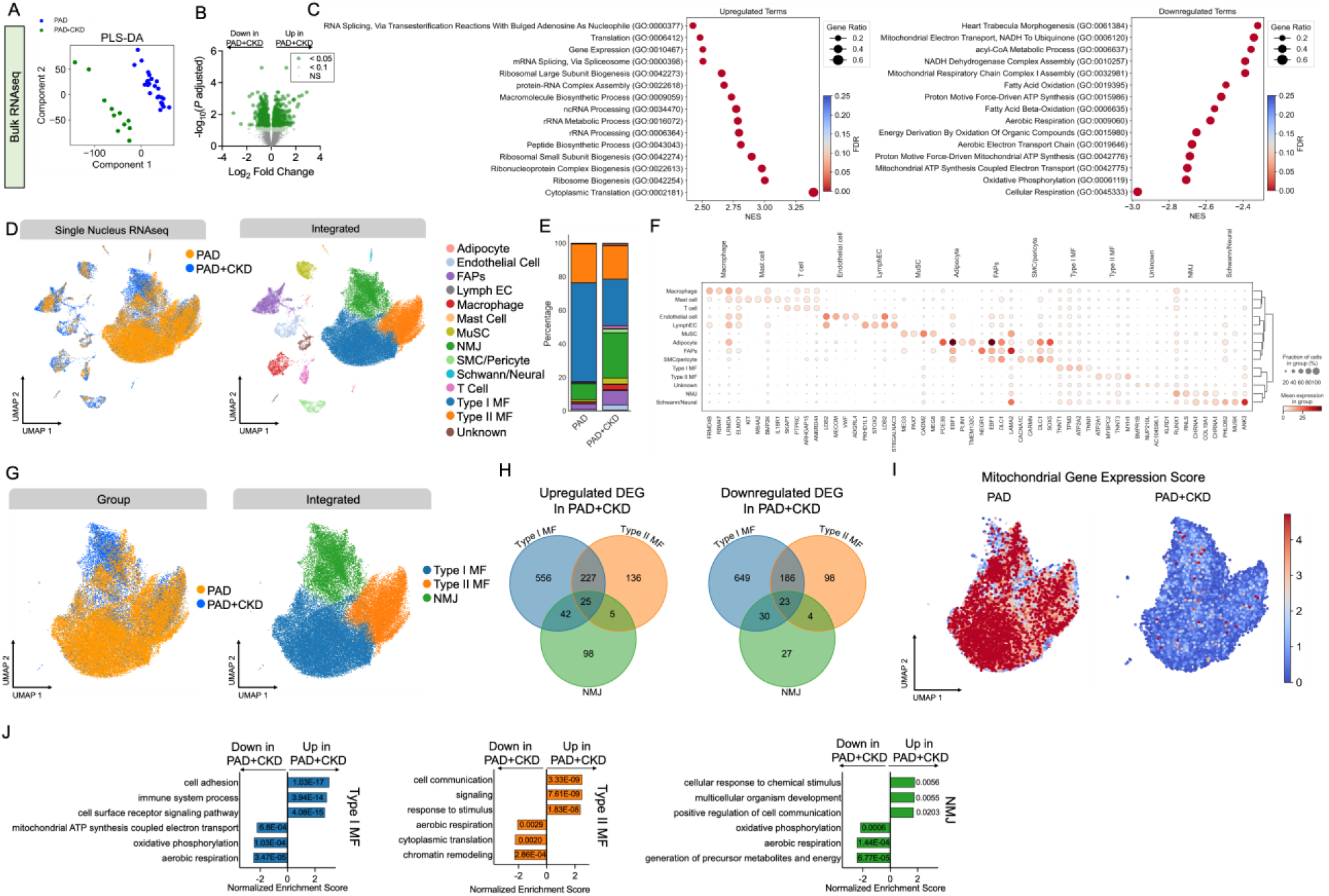
Whole muscle and single nucleus RNA sequencing identify mitochondrial deficiency and cytoplasmic translation defects in patients with PAD+CKD. (**A**) RNA sequencing on total RNA from the gastrocnemius was performed (n=88 patients). Partial least squares discriminant analysis (PLS-DA) revealed clear separation between PAD and PAD+CKD patients. (**B**) Volcano plot of gene expression shows differentially expressed genes in PAD and PAD+CKD patients. (**C**) Gene set enrichment analysis in significantly upregulated and downregulated genes between PAD and PAD+CKD patients. (**D**) Single nucleus RNA sequencing was performed on gastrocnemius muscle specimens from 20 PAD and 12 PAD+CKD patients. UMAPs presented by group and by cell types are shown. (**E**) Percentage of nuclei within each cluster determined by group. (**F**) Dotplots of the top 4 marker genes for each cell type. (**G**) UMAPs of the subclustering of myofiber nuclei by group and type. (**H**) Venn diagrams shown differentially expressed genes by myonuclei type. (**I**) UMAPs showing the mitochondrial gene module score for each nuclei demonstrated the mitochondrial deficiency in PAD+CKD muscles. (**J**) Gene set enrichment analysis results for myonuclei populations. The x-axis is the normalized enrichment score and the values for each bar are the adjusted *P*-values.

Because there are many cell types within skeletal muscle, we next used single nucleus RNA sequencing to enhance the resolution of analysis of calf muscle. 61,586 nuclei were analyzed from 20 PAD patients and 12 patients with PAD+CKD. Unsupervised clustering identified 14 clusters representing the major cell types expected in skeletal muscle (**Figure 5D**). Compared to PAD patients without CKD, PAD+CKD patients had fewer Type I myofiber nuclei and a higher proportion of nuclei that represented NMJ/regenerating myonuclei, fibro-adipogenic progenitor cells (FAPs), endothelial cells, and macrophages (**Figure 5E**). Top marker genes for each cell/nuclei type are shown in **Figure 5F**. DEG analysis in the myofiber nuclei populations (**Figure 5G,H**) revealed a significant number of differentially expressed genes within each cluster, however Type I myofiber nuclei had the largest number of DEGs. With knowledge that bulk RNAseq GSEA analysis identified mitochondria as a downregulated biological process, we generated a mitochondrial gene expression score which represents the overall mitochondrial gene expression. This analysis confirmed all myonuclei populations in PAD+CKD muscles displayed a mitochondrial deficiency, although Type I myofiber nuclei appeared to be most affected (**Figure 5I**). GSEA analysis from myonuclei had common pathways related to “aerobic respiration” in the downregulated DEGs (**Figure 5J**). Additional DEG, GSEA, and top marker genes for other cell types can be found in Supplemental Dataset 4.

### Fibro-adipogenic progenitor (FAPs) cells are uniquely remodeled in muscle from patients with PAD and CKD

Next, we investigated how CKD impact FAPs, a resident mesenchymal stem cell population that has been shown to be critical for muscle regeneration, were impacted by CKD in patients with PAD. Subclustering of FAPs revealed eight distinct cell populations including pro-fibrotic, adipogenic, and MME+ clusters^32^ (**Figure 6A**). PAD patients with CKD had a higher abundance of adipocyte/adipogenic FAPs (16% vs. 5.6%) and a unique population of ADAM12^+^ FAPs that was only present in PAD+CKD (**Figure 6B**). We used the Palantir algorithm^33^ to characterize differences in cell fate between PAD and PAD+CKD FAPs. In patients with only PAD, a single terminate state representing the low abundance adipocyte lineage was observed, whereas two clear terminal states were seen in PAD+CKD FAPs (adipocyte and MME^+^) along with a substantially higher differentiation potential in other populations compared with FAPs from PAD patients (**Figure 6C**). Trajectory inference uncovered and additional pathway for FAP differentiation in patients with PAD+CKD producing the ADAM12^+^ population (**Figure 6D**). Normalized gene expression showed the ADAM12^+^ FAP population was enriched for myokines IL6 and IGF1, whereas the pro-fibrotic FAPs displayed high expression of TGFB1 and COL1A1 (**Figure 6E**). Expression of branch-specific genes along pseudotime are shown in **Figure 6F**. Other notable cell types that displayed significant transcriptome and population differences in PAD+CKD included macrophages (**Supplemental Figure 3**) and endothelial cells (**Supplemental Figure 4**).

**Figure 6.**
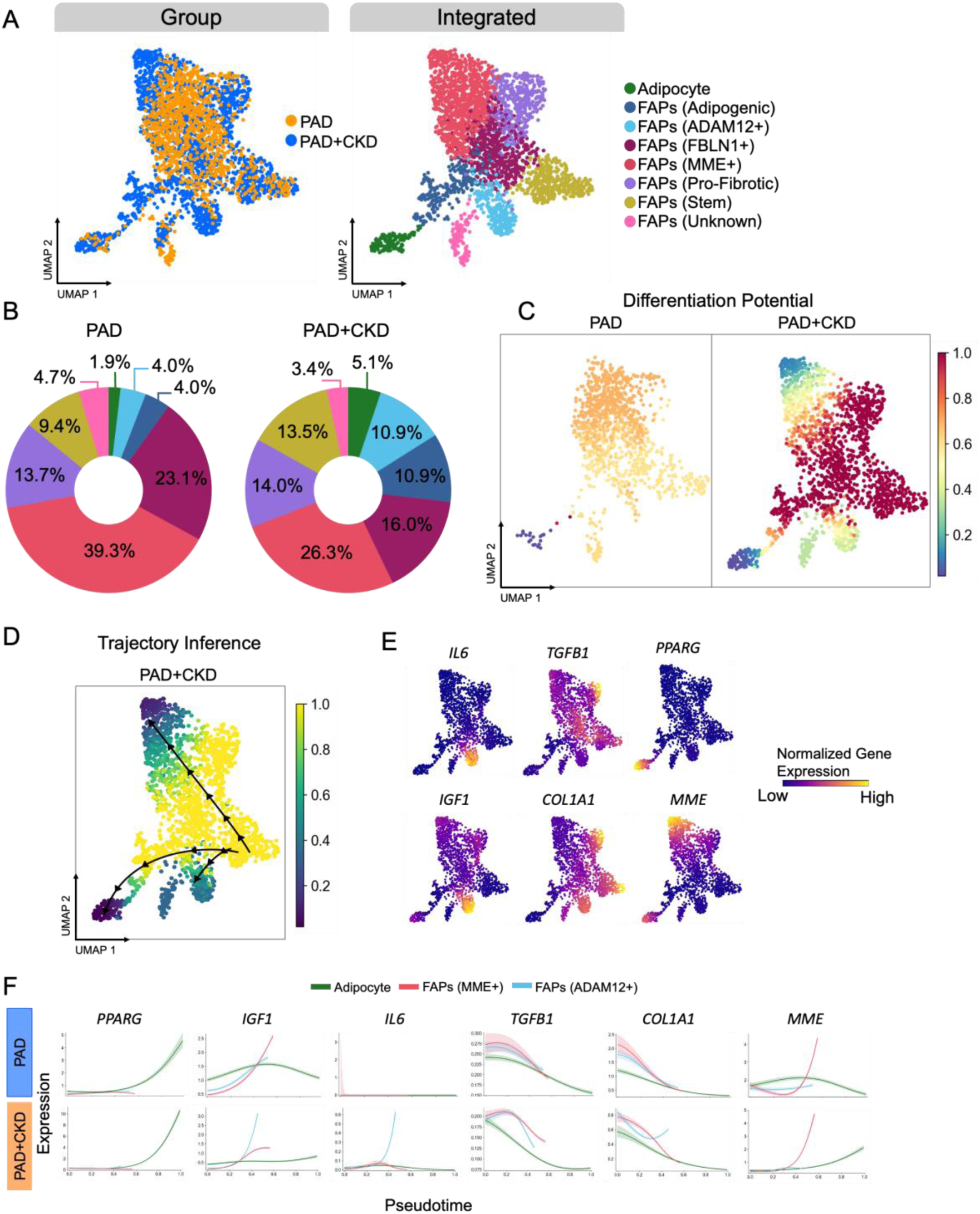
Fibro-adipogenic progenitor cells are uniquely remodeled in muscles from patients with PAD+CKD. (**A**) UMAPs of the subclustering of FAPs presented by group and by subpopulation type are shown. (**B**) Donut plots showing the abundance of subpopulations of FAPs in both groups. (**C**) Ranked significant ligand-receptor communications for relative information flow between AHR^fl/fl^ and AHR^mKO^ muscles. (**D**) Trajectory inference in FAPs showed three distinct fates in PAD+CKD. (**E**) Feature plots showing normalized gene expression levels for inflammatory, pro-fibrotic, and adipogenic genes. (**F**) Gene expression changes across those three fates across pseudotime for both groups.

### CKD influences intercellular communication in calf muscle from patients with PAD

To explore how the presence of CKD influences intercellular communication, we employed CellChat^34^ to assess inferred ligand-receptor interactions. Circle plots depicting the global intercellular communication are shown in **Figure 7A**, where the line width represents the communication strength. While the number of interactions was lower in PAD+CKD compared to PAD (2920 vs. 3552), the strength of those interactions was greater (**Figure 7B**). Comparisons of the major ‘senders’ and ‘receivers’ identified FAPs and adipocytes as the primary ‘senders’ whereas vascular populations including endothelial, lymphatic endothelial, and smooth muscle/pericyte cells were determined to be the major ‘receivers’ (**Figure 7C**). Significant signaling pathways between PAD and PAD+CKD were identified according to variations in information flow within inferred signaling networks (**Figure 7D**)

**Figure 7.**
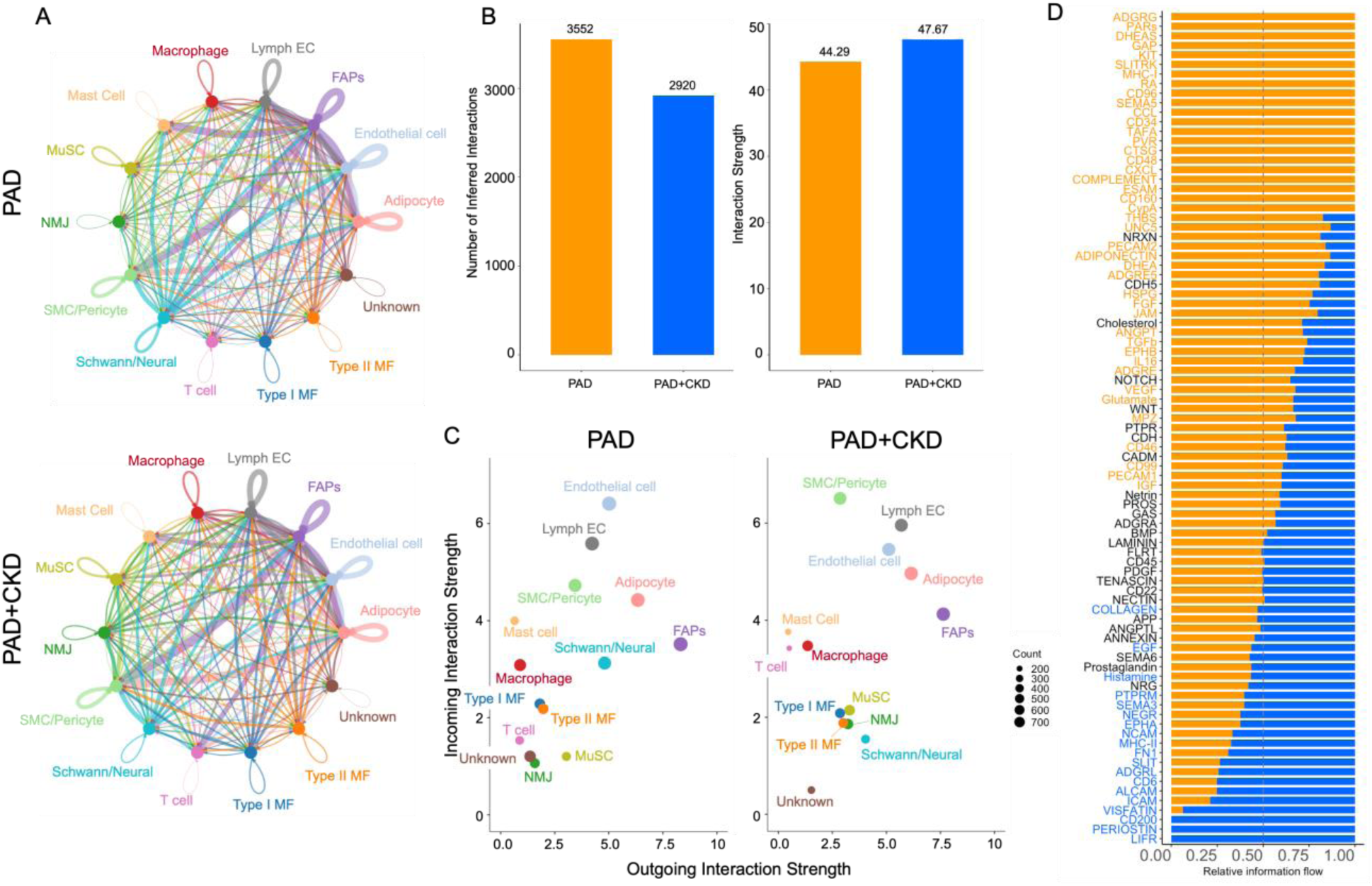
CellChat analysis predicts changes in intercellular communication of PAD+CKD patients. (**A**) Circle plots showing the overall intercellular communication occurring in non-PAD and PAD. Circle sizes represent the number of cells and edge width represents communication probability. (**B**) Comparison of outgoing and incoming interaction strengths for all cells type in non-PAD and PAD. (**C**) Ranked significant ligand-receptor communications for relative information flow between non-PAD and PAD muscles. (**D**) Circle plots for FGF signaling communication in non-PAD and PAD.

## DISCUSSION

Patients with PAD often present with one or more comorbid conditions that worsen ischemic pathology and increase the risk of an adverse limb event. Recent studies have clearly demonstrated that CKD may be one of the strongest risk factors for poor outcomes in PAD^11,35^. In this cross-sectional study, we leveraged state-of-the-art multiomic analyses and mitochondrial and muscle function assessments to dissect how CKD impacts calf muscle pathophysiology in patients with PAD. Compared to non-PAD control with normal kidney function, patients with only PAD, patients with only CKD, and patients with both PAD and CKD had significantly weaker calf muscles and muscle fiber areas (Figure 1). Compared to patients with PAD, those with PAD and CKD were also significantly weaker and had smaller muscle fiber areas demonstrating that CKD exacerbates the ischemic myopathy in PAD. Skeletal muscle mitochondrial function was significantly impaired in patients with CKD only and those with PAD and CKD (Figure 2). Interestingly, no differences were observed between patients with PAD and the non-PAD controls. A significant negative correlation was found between the levels of indoxyl sulfate, L-Kynurenine, or the L-Kyn/Trp ratio and both mitochondrial function and muscle strength (Figure 4). This observation agrees with previous studies that reported similar associations in rodents with and without CKD ^23,28–30^, implicating uremic toxins as strong drivers of muscle impairment in CKD. These findings also agree with those of preclinical studies in which limb function of animals with CKD subjected to hindlimb ischemia was impaired compared to animals with normal kidney function subjected to hindlimb ischemia ^11,30,36–38^. Mechanistically, recent studies have shown that the aryl hydrocarbon receptor is required for tryptophan-derived uremic toxins to impair skeletal muscle function and ischemic angiogenesis^11,30,38^. Beyond uremic toxins, tobacco smoking products also contain ligands that activate the aryl hydrocarbon receptor in skeletal muscle^39,40^, namely 2,3,7,8-Tetrachlorodibenzodioxin (known as TCDD), and smoking is considered the strongest risk factor for PAD suggesting this pathway could have a much broader impact of calf muscle pathophysiology in patients with PAD. In totality, these findings suggest that CKD induces muscle mitochondrial dysfunction in patients with and without PAD, potentially through mechanisms involving uremic toxins like indoxyl sulfate and kynurenines that activate the aryl hydrocarbon receptor.

Calf muscle mitochondrial OXPHOS was ∼40% lower in patients with CKD than those without (Figure 2). This profound impact of CKD on skeletal muscle mitochondrial function and content aligns with previous studies^21,24,41–43^. In contrast, there were no differences between non-PAD controls and patients with PAD that had normal kidney function. This lack of difference in mitochondrial respiratory function is consistent with some previous studies^44–46^, but not all^47–50^, that used similar methodologies. The lack of agreement across these studies may be due to modest sample sizes and differences in patient characteristics. Nonetheless, there is accumulating evidence that markers of mitochondrial content (mtDNA copy number, electron transport system complex abundance, citrate synthase activity, etc.) are increased in the calf muscle of patients with PAD^17,51,52^. This compensatory response is likely related to the hypoxia-induced energetic stress response that occurs within the calf muscles of PAD^17^. The striking impact of CKD on calf muscle mitochondrial health was also supported by the down regulation of mitochondrial gene expression observed in both bulk and single nucleus RNA sequencing analyses comparing PAD and PAD+CKD groups (Figure 5). In both diseases calf muscle mitochondrial function correlates with walking performance^24,52,53^. Further investigation is needed to determine whether interventions that improve or optimize calf muscle mitochondrial function can lead to improvements in walking performance in these patients.

Calf muscle from patients with PAD and CKD displayed a unique remodeling of a resident mesenchymal stem population called fibro-adipogenic progenitor cells (FAPs). These resident multipotent stem cells are located within the muscle interstitium where they have a heavy presence surrounding blood vessels. They have numerous differentiation paths with the two main fates being adipocytes or myofibroblasts^54^. Compared to PAD patients with normal kidney function, those with PAD and CKD had ∼2.5-fold increase in adipocytes according to snRNAseq results. This accumulation of intramuscular adipose tissue is a form of myosteatosis which has been shown to impair muscle function^55,56^ and regeneration from injury^57–59^. This phenotype has been observed independently in CKD^60^ and PAD^61^. Considering that muscle function is strongly associated with mortality in PAD^13,15^ and CKD^62^, further studies are needed to determine if therapies the prevent or mitigate intramuscular adipose tissue in these conditions can improve muscle function and lower mortality risk.

Plasma metabolomics analyses revealed the wide-ranging impact on the plasma metabolome in patients with CKD (>500 differentially abundant biochemicals). In contrast, we found relatively few metabolites (99) that were differentially abundant between non-PAD controls and PAD patients. Only a few studies have examined the plasma metabolomes in patients with PAD^11,63–65^, with the majority of studies reporting associations between amino acid metabolites, namely kynurenine and tryptophan, as well as indoles, and the risk of developing PAD or having adverse limb events^11,64,65^. The link between these uremic toxins and PAD risk may be indicative of the incidence of renal dysfunction in those study populations despite some studies excluding patients with diagnosed kidney disease. Alternatively, the source of kynurenines and indoles could be related to levels of systemic inflammation or dysregulation of the gut microbiome^66–68^, both areas that have been underexplored in PAD. The highly overlapping plasma metabolomes between the non-PAD controls and patients with PAD may also be indicative of how well these groups were matched for co-morbid conditions, risk factors, and medication usage (Table 1). Additionally, the plasma metabolome in PAD is not likely to match the complex microenvironment within the affected limb where local metabolite alterations are presumed to be much greater than those reflected from a systemic plasma sample. Local sample collection from the diseased limb followed by subsequent metabolomic analyses could reveal the full impact of limb ischemia on metabolite exchange in tissues below of the occlusive lesion.

Given that our cohort of patients were restricted to moderate CKD (Stage 3b-4) and moderate-severe PAD (Rutherford Stage 3-5), our analyses may not be representative of the full spectrum of either disease. However, this design was chosen to enable deep phenotyping and multiomic analyses across the four groups. Additional studies should be performed to evaluate the reported relationships across the spectrum of both PAD and CKD. Although this patient cohort contained 39% female participants, we have not performed sex comparisons due to the relatively small sample sizes within each group for such a comparison. Future analyses of these data could be performed to begin understanding how biological sex may alter the impact of CKD on PAD pathobiology. Additionally, while the control group was relatively well matched for clinical and physical characteristics, there were some comorbid conditions that were more prevalent in the PAD and CKD groups including hypertension and diabetes. It is important to note that these two conditions are among the strongest risk factors for developing CKD. This was a cross-sectional study design and longitudinal analyses were not performed. Thus, analyses exploring how specific outcomes change across time and whether they impact adverse limb events, morbidity, or mortality risk could not be performed. Despite these limitations, this study provides a robust and in-depth analysis of the skeletal myopathy in patients with PAD and CKD using physiological testing and multiomic analyses which identify pathways and processes that may have potential for interventional approaches that have promise to improve muscle function in these patients.

## CONCLUSIONS

This study demonstrates that chronic kidney disease significantly worsens the ischemic limb myopathy in patients with PAD evidenced by lower calf muscle strength, muscle fiber sizes, and mitochondrial function. Multiomic analyses of calf muscle revealed a striking deficiency in myonuclear gene expression related to mitochondria and protein translation specifically in patients with both PAD and CKD, compared to those with only PAD. The significant correlation between plasma uremic toxin levels and both calf muscle strength and mitochondrial function suggests that therapeutic strategies aimed at reducing uremic toxins may hold promise for improving muscle health in patients with CKD.

## Data Availability

The data supporting the conclusions of this study are available from the corresponding author.

## Sources of Funding

This study was supported by National Institutes of Health (NIH) grants R01-HL149704 and HL171050 (T.E.R.). S.T.S. was supported by NIH grant R01HL148597. S.A.B. was supported by NIH grant R01DK119274. K.K. was supported by the American Heart Association grant POST903198. T.T. was supported by NIH grant F31-DK128920. V.R.P. was supported by the American Heart Association grant 24PRE1193999. C.P. was supported by the American Heart Association grant 24PRE1196311.

## Disclosures

None.

## Supplemental Material

Expanded Materials and Methods

Supplemental Figures 1-4

Supplemental Dataset 1 (metabolomics).XLSX

Supplemental Dataset 2 (metabolomics pathway).XLSX

Supplemental Dataset 3 (bulk RNA).xlsx

Supplemental Dataset 4 (snRNA).xlsx

## ABBREVIATIONS

CKD: chronic kidney disease
GFR: glomerular filtration rate
GSEA: gene set enrichment analysis
OXPHOS: oxidative phosphorylation
PAD: peripheral arterial disease
ROS: reactive oxygen species
snRNAseq: single nucleus RNA sequencing

## NOVELTY AND SIGNIFICANCE

### What is known?

- Peripheral artery disease (PAD) patients with chronic kidney disease (CKD) have poorer health outcomes and increased risk for adverse limb events and mortality compared with patients with normal kidney function.
- The accumulation of uremic toxins in CKD have been associated with adverse limb events in patients that have PAD.

### What new information does this article contribute?

- Patients with PAD and CKD have significantly lower calf muscle strength, muscle fiber size, and mitochondrial function compared to PAD patients with normal kidney function.
- CKD, but not PAD alone, has a significant impact on the plasma metabolome.
- Uremic toxins such as indoxyl sulfate and L-Kynurenine have significant inverse relationships with calf muscle strength and mitochondrial function.
- Bulk and single nucleus RNA sequencing reveals a mitochondrial deficiency in myofibers and pro-fibrotic/pro-adipogenic differentiation states of fibro-adipogenic progenitor cells in patients with PAD and CKD.

There is overwhelming evidence that PAD patients with CKD have significantly higher risk of major adverse limb events and death compared to PAD patients without CKD. Moreover, having CKD increases the likelihood that endovascular and open revascularization procedures fail in patients with PAD. Skeletal muscle pathologies have been shown to play important roles in both PAD and CKD independently. Because skeletal muscle function is a strong predictor of morbidity and mortality, it is important to understand how CKD exacerbates the myopathy of PAD to improve medical management in these patients. In a cohort of 123 patients, calf muscle strength was lowest in PAD patients with CKD, although both those PAD and CKD alone were also significantly weaker than non-PAD and non-CKD controls. Muscle fiber atrophy was also evident and muscle fiber size correlated with calf muscle strength. Using calf muscle biopsies, we found that mitochondrial function was significantly impaired in patients with CKD and those with PAD+CKD. Significant inverse correlations were found between uremic toxins and muscle strength and mitochondrial function. Bulk and single nucleus RNA sequencing revealed the wide-ranging impact of CKD on the muscle transcriptome in patients with PAD. In summary, this study establishes CKD as a major driver of skeletal muscle pathology in patients with PAD.

## Notes

### Competing Interest Statement

The authors have declared no competing interest.

### Clinical Trial

This was not a clinical trial.

### Author Declarations

This study was approved by the institutional review boards at the University of Florida and the Malcom Randall VA Medical Center (Protocol IRB201801553).

